# The relationship between visually evoked effects and concussion in youth

**DOI:** 10.1101/2021.12.03.21267248

**Authors:** Carlyn Patterson Gentile, Geoffrey K Aguirre, Kristy B. Arbogast, Christina L. Master

**Author notes:** **<u>Commercial Relationships Disclosures:</u>** C.P.G.: none; G.K.A.: none; K.B.A.: none; C.L.M.: none.

## Abstract

Increased sensitivity to light is common following concussion. Viewing a flickering light can also produce uncomfortable somatic sensations like nausea or headache. Here we examined effects evoked by viewing a patterned, flickering screen in a cohort of 81 uninjured youth athletes and 84 youth with concussion. We used exploratory factor analysis and identified two primary dimensions of variation: the presence or absence of visually evoked effects, and variation in the tendency to manifest effects that localized to the eyes (e.g., eye watering), versus more generalized neurologic symptoms (e.g., headache). Based on these two primary dimensions, we grouped participants into three categories of evoked symptomatology: no effects, eye-predominant effects, and brain-predominant effects. A similar proportion of participants reported eye-predominant effects in the uninjured (33.3%) and concussion (32.1%) groups. By contrast, participants who experienced brain-predominant effects were almost entirely from the concussion group (1.2% of uninjured, 35.7% of concussed). The presence of brain-predominant effects was associated with a higher concussion symptom burden and reduced performance on visio-vestibular tasks. Our findings indicate that the experience of negative constitutional, somatic sensations in response to a dynamic visual stimulus is a salient marker of concussion and is indicative of more severe concussion symptomatology. We speculate that differences in visually evoked effects reflect varying levels of activation of the trigeminal nociceptive system.

## INTRODUCTION

Visual symptoms and eye movement abnormalities are common following head trauma^1^ and have been explored as physiologic biomarkers for concussion. As an example, dynamic pupillary responses are increased in children following concussion, and have been proposed as an objective biomarker^2,3^. Similarly, deficiencies in saccadic eye movements, vestibulo-ocular reflex (VOR), and visual motion sensitivity (VMS) are all associated with concussion^4–7^.

It is also well-established that light sensitivity can be enhanced following concussion^8^ and thus, is a standard question on concussion symptom inventories^9–11^. Visually evoked effects offer an intriguing avenue for concussion assessment – including evaluations that extend beyond effects localized to the eye. High contrast patterned and flickering light is uncomfortable to view and can induce multiple somatic sensations^12^, however the somatic effects evoked by intense visual stimuli have not been assessed as a marker for concussion. In migraine—for which light sensitivity is a cardinal symptom—visual discomfort and somatic sensations from flickering light is a common inter-ictal complaint^13,14^. These stimuli are also associated with enhanced activity in the visual cortex in migraine^15,16^ and photophobia acutely following concussion^17^. The symptoms evoked by patterned flickering light may serve as a biomarker and provide a window into the neural pathways involved in concussion.

The purpose of this investigation was to determine if a patterned flickering visual stimulus discriminated between uninjured athletes and concussed youth based on the characteristics of the signs and symptoms provoked.

## METHODS

### Participants

This study is a retrospective analysis of data collected from a prospective enrolled cohort. We studied youth between 13 and 20 years old participating in a broader study on youth concussion through the Children’s Hospital of Philadelphia (CHOP) Minds Matter Concussion Program from February 2018 to October 2021. Uninjured participants were recruited through the sports teams of a local Philadelphia-area school. They were offered an opportunity to participate in clinical assessments at the beginning of and immediately after their sports season. Only the first session they participated in was used in this analysis. For the concussion group, participants were recruited from both the CHOP Minds Matter Concussion Program, as well as the high school where healthy youth were enrolled. Diagnosis of concussion was determined by a trained sports medicine pediatrician according to the Consensus Statement on Concussion in Sports^18^. All concussed participants’ data sessions were within 28 days of their concussion. Assent and consent were obtained from participants and guardians, respectively, and the study was approved by the CHOP Institutional Review Board in accordance with the Declaration of Helsinki. Participants were prescreened to ensure normal or corrected to normal visual acuity under binocular and monocular viewing conditions using a Snellen visual acuity chart at 10 feet. Fifty-nine participants (20 uninjured, 39 concussed) wore corrective lenses during testing. Age, sex, race, ethnicity, and concussion and migraine history (self and family) were self-reported for uninjured participants and abstracted from the medical record for concussed participants.

### Data collection sessions

Data were collected from uninjured youth during a single recording session as part of a pre- or post-season sports evaluation. Data were collected from concussed youth at a single recording session during clinical visits for concussion management. In the case of a participant having multiple clinical visits within the first 28 days of their concussion, only the first visit assessing visually evoked effects was used. Trained research staff conducted clinical assessments in either the athletic training room at the high school, primarily for uninjured athletes, or the sports medicine office, primarily for concussed participants.

#### Symptom inventory

All participants completed the Post-Concussion Symptom Inventory (PCSI), a scale validated in youth for measuring concussion symptoms^19^. The question ‘In general, to what degree do you feel “differently” than before the injury (not feeling like yourself)?’ was excluded from the total PCSI score for comparison across uninjured and concussed groups as this question was not applicable to the uninjured group.

#### Visio-vestibular assessment

All participants underwent visio-vestibular examination (VVE), a battery of physical exam metrics that assess visual and vestibular function. These elements included 1) smooth pursuit for 5 repetitions, 2) horizontal and vertical saccades for 30 repetitions, 3) horizontal and vertical vestibulo-ocular reflex (VOR) for 30 repetitions, 4) Visual motion sensitivity (VMS) for 5 repetitions, and 5) near point convergence, with abnormal defined as a break point where vision becomes double as greater than 6cm. Abnormality for smooth pursuits, saccades, VOR, and VMS was defined as provocation of symptoms within the number of repetitions that required the participant to stop testing. For more detailed information on testing refer to our prior work, which has shown the testing to be reliable and specific for concussion across multiple clinical settings^7,20–23^. Participants with symptom exacerbation by definition cannot complete the target number of repetitions.

#### Visual flicker

Participants viewed a wide field 85% contrast checkerboard with a pattern reversal rate of 2 reversals per second. Stimuli were presented for a continuous 20 seconds for a total of 40 reversals in a single block. The block was repeated 5 times in a single session. Because these data were being collected along with visual evoked potentials, visual stimuli largely reflected ISCEV recommendations for visual evoked potentials^24^. The full description of the visual stimulus conditions can be found in our prior report^25^.

#### Visually evoked effects

Eye movements were assessed during and after each recording session. While examining eye movements, experimenters also documented the presence or absence of the following physical signs of visual discomfort: eyes watering, eyes reddening, eyes slowing, circular eye movements, or “other signs”. At the end of the recording session participants were asked if they experienced provocation or worsening of the following symptoms: dizziness, headache, nausea, eye pain, eye fatigue, or other symptoms. No participants displayed circular eye movements, eye slowing, and no other signs were reported, so these were not included in data analysis. Other symptoms which were entered via free text included ‘eyes tired’ (4), ‘eyes foggy’ (1), ‘eye strain’ (1), ‘blurry vision’ ‘blurriness’ or ‘eye blur’ (2), ‘double vision’ (1), ‘eyes dry’ (2), ‘feeling out of it’ (1). Because the ‘other symptoms’ responses were heterogenous, and the same symptoms were not frequently reported, these symptoms were excluded from data analysis.

### Data Analysis

All analyses were performed using custom written code in Matlab (Mathworks, Natick, MA). Code is publicly available: https://github.com/pattersongentilelab/visuallyEvokedEffects.

#### Demographic data

Demographics were compared between the uninjured and concussion groups using a *χ*^2^ test. P-value < 0.05 was used as the threshold for significance throughout. Concussion history, migraine history, and migraine family history were also compared, as some studies^26,27^ (although not all^28^), indicate these are predictors for prolonged concussion symptoms. The median was reported for age (in years), days post-injury, and individual and total PCSI scores. All values were reported with 95% confidence intervals derived by bootstrap analysis.

#### Analysis of visually evoked effects

Multiple correspondence analysis (MCA) was used across all participants to reduce the dimensionality of 8 evoked signs and symptoms: dizziness, headache, nausea, eyes watering, eyes tearing, eye fatigue, eyes reddening, and eye pain. The code used to perform MCA was based on the indicator matrix, adapted from publicly available code^29^. The first two dimensions represented (1) the presence or absence of visually evoked symptoms, and (2) whether signs and symptoms were eye-predominant or brain-predominant. These two primary dimensions were used to separate participants into 3 visually evoked effects categories: (1) no effects, defined as a dimension 1 coefficient of -1 or less, (2) eye-predominant effects defined as a dimension 1 coefficient of greater than -1 and dimension 2 coefficient of 0.1 or less, and (3) brain-predominant effects defined as a dimension 1 coefficient greater than -1 and a dimension 2 coefficient of greater than 0.1.

The percentage of participants per visually evoked effects category was compared between uninjured and concussion groups using the Kruskal-Wallis test. Due to large differences in symptom burden between concussion and uninjured participants as measured by total PCSI score, and unequal representation of concussion and uninjured participants across the three groups, statistical analysis for differences between visually evoked effects categories was conducted on the uninjured and concussed groups separately. Kruskal-Wallis was used to compare visually evoked effects categories across participant demographics, PCSI scores, and *χ*^2^, and p-values were reported. Multiple comparisons with Tukey method were used to determine significance between individual visually evoked effects categories. Performance on the number of repetitions of VVE metrics was compared between groups using a two-sided, two-way ANOVA with visually evoked effects group and uninjured vs. concussion groups as predictors. Multiple comparisons with Tukey method were also used for this analysis to determine significance between effects categories.

## RESULTS

Data from 81 uninjured participants and 84 concussed participants were included in analysis. Uninjured and concussed groups did not differ significantly in age, biological sex, or racial/ethnic identity (Table 1). Our cohort was overall similar to the demographics of the state of Pennsylvania^30^, with the exception of an under-representation of participants who identify as Hispanic. Compared to uninjured youth, a greater percentage of concussed youth reported history of prior concussion (27.1% vs. 44.0%; *χ*^2^ = 5.09, p = 0.02) and family history of migraine (9.9% vs. 34.5%; *χ*^2^ = 12.61, p = 3.8e^-4^).

**Table 1:**
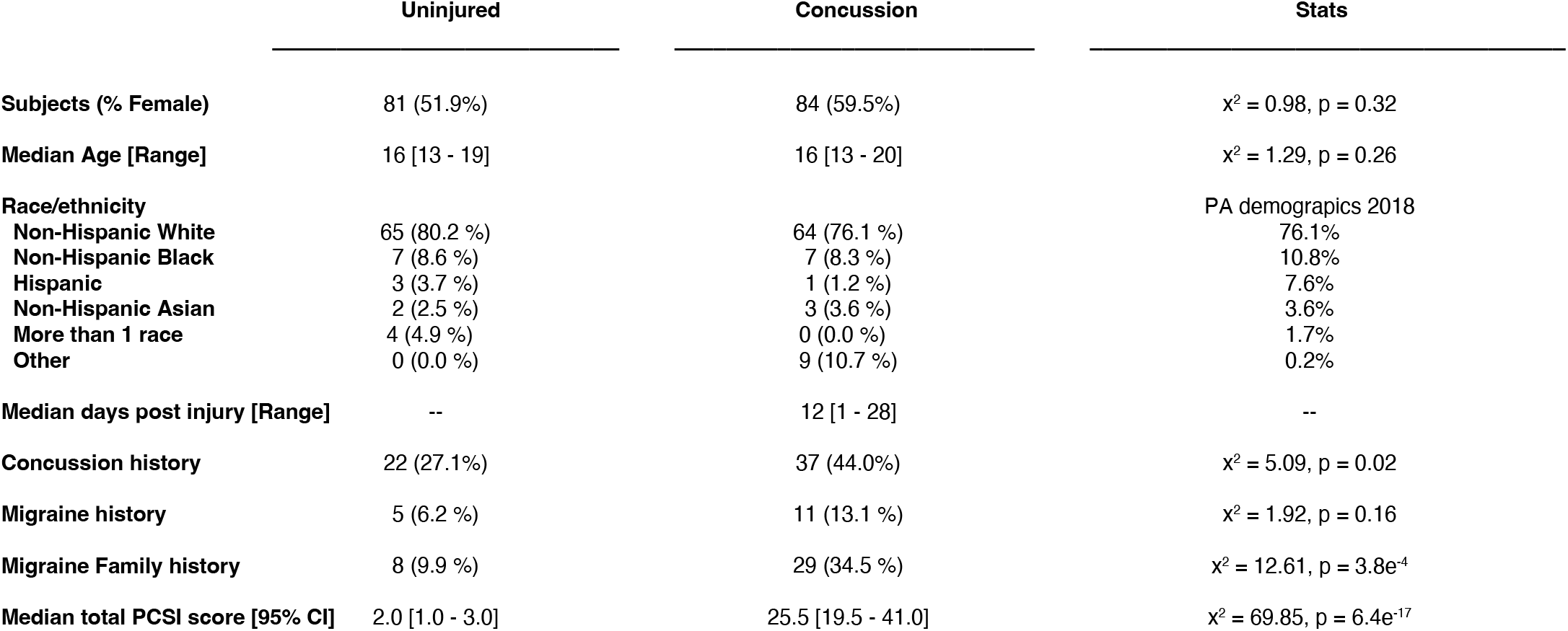
uninjured vs. concussion group demographics.

|  | Uninjured | Concussion | Stats |
| --- | --- | --- | --- |
| **Subjects (% Female)** | 81 (51.9%) | 84 (59.5%) | $\chi^2 = 0.98$, $p = 0.32$ |
| **Median Age [Range]** | 16 [13 - 19] | 16 [13 - 20] | $\chi^2 = 1.29$, $p = 0.26$ |
| **Race/ethnicity** |  |  | PA demographics 2018 |
| **Non-Hispanic White** | 65 (80.2 %) | 64 (76.1 %) | 76.1% |
| **Non-Hispanic Black** | 7 (8.6 %) | 7 (8.3 %) | 10.8% |
| **Hispanic** | 3 (3.7 %) | 1 (1.2 %) | 7.6% |
| **Non-Hispanic Asian** | 2 (2.5 %) | 3 (3.6 %) | 3.6% |
| **More than 1 race** | 4 (4.9 %) | 0 (0.0 %) | 1.7% |
| **Other** | 0 (0.0 %) | 9 (10.7 %) | 0.2% |
| **Median days post injury [Range]** | -- | 12 [1 - 28] | -- |
| **Concussion history** | 22 (27.1%) | 37 (44.0%) | $\chi^2 = 5.09$, $p = 0.02$ |
| **Migraine history** | 5 (6.2 %) | 11 (13.1 %) | $\chi^2 = 1.92$, $p = 0.16$ |
| **Migraine Family history** | 8 (9.9 %) | 29 (34.5 %) | $\chi^2 = 12.61$, $p = 3.8e^{-4}$ |
| **Median total PCSI score [95% CI]** | 2.0 [1.0 - 3.0] | 25.5 [19.5 - 41.0] | $\chi^2 = 69.85$, $p = 6.4e^{-17}$ |

### Visually evoked effects

MCA of 8 visually evoked effects was performed to identify underlying structures within the data. Close to half of the variability (44.7%) across participants was explained by the first two dimensions (Figure 1a). The first dimension, which accounted for 26.5% of the variability, represents the absence (negative direction) or presence (positive direction) of visually evoked effects. Concussed participants had a higher mean dimension 1 coefficient compared to uninjured participants: 0.78 [95%CI: 0.33, 1.21] vs. -0.82 [95%CI: -1.03, -0.55] indicating concussed participants reported more visually evoked effects than uninjured participants.

**Figure 1:**
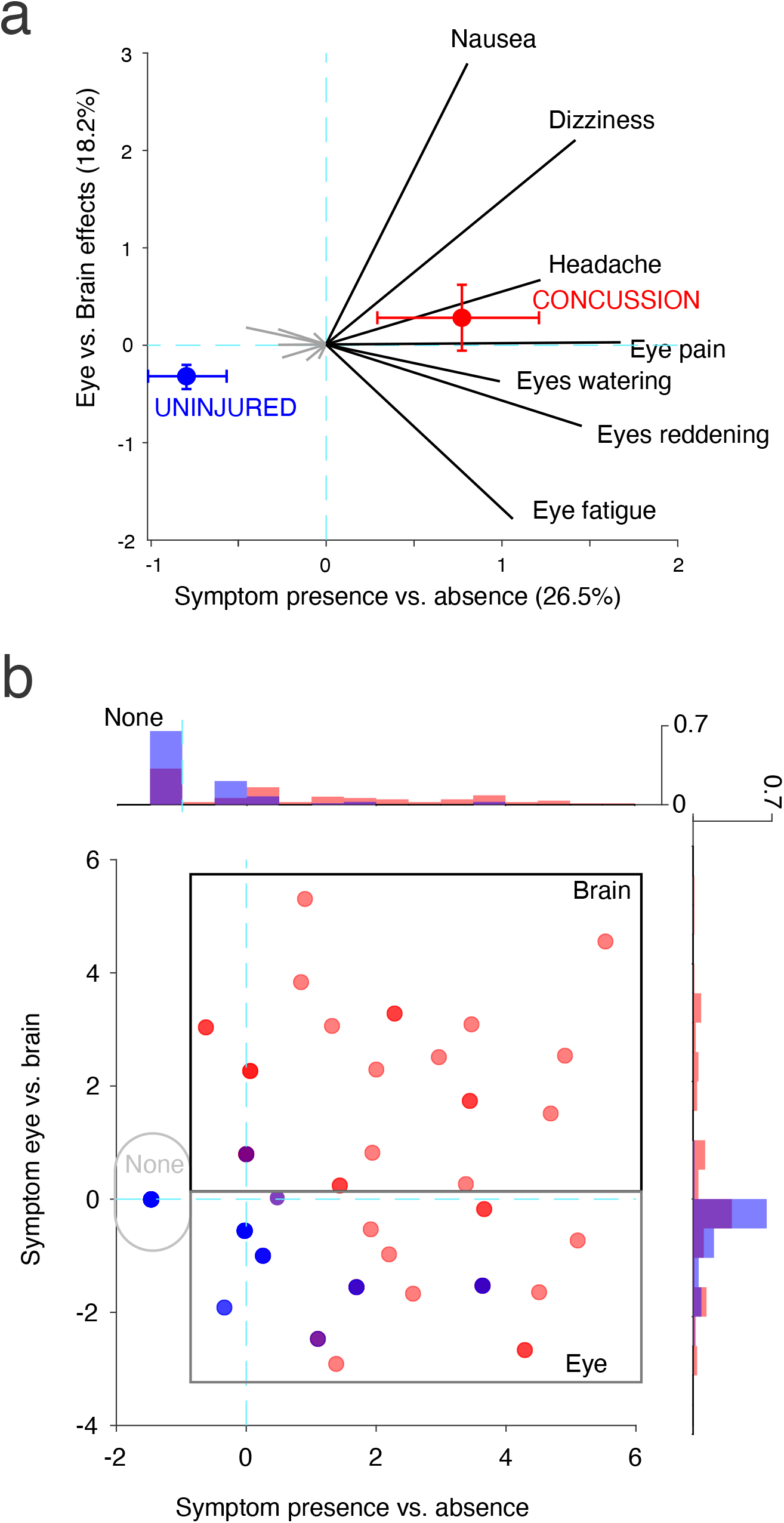
MCA for visually evoked effects.

The second dimension, which accounted for 18.2% of the variability, split effects qualitatively. Dizziness, headache, and nausea were associated with a larger positive coefficient, while eye fatigue, eyes watering, eyes reddening, and eye pain were associated with a smaller or negative coefficient. We adopt here the short-hand terms “eye effects” (negative values) and “brain effects” (positive values) to describe the two ends of variation along this second dimension revealed by the MCA. Concussed participants had a higher mean dimension 2 score compared to uninjured participants: 0.29 [95%CI -0.07, 0.64] suggesting that the concussed participants overall reported more brain effects. Uninjured participants had a negative mean coefficient of - 0.30 [95%CI -0.43, -0.19] suggesting that when visually evoked effects were present, they were more likely to be eye effects.

### Brain-predominant visually evoked effects are associated with concussion and higher symptom burden

MCA dimension 1 and dimension 2 coefficients were used to categorize participants into three categories: those who experienced (1) no effects, (2) eye-predominant effects, or (3) brain-predominant effects (Figure 1b). There was some heterogeneity across concussed participants in these groups: 27% of those in the eye-predominant group also reported headache, and 50% of the brain-predominant group reported at least one eye effect (Table S1).

The percentage of uninjured and concussed participants within each effect category differed (Figure 2a). Most uninjured participants reported no effects (65.4%), with fewer reporting eye-predominant effects (33.3%), and only one participant reporting brain-predominant effects (1.2%). By comparison, there were similar percentages of concussed participants within each of the 3 groups: 32.1% reported no effects, 32.1% reported eye-predominant effects, and 35.7% reported brain-predominant effects (uninjured v. concussion *χ*^2^ = 35.32, p = 2.1e^-6^). There was a significant effect of visually evoked effects category on total PCSI score for concussed participants (*χ*^2^ = 15.39, p = 4.5e^-4^). Median PCSI score [95%CI] of concussed participants was 9.0 [7.0, 13.0] for no effects, 33.0 [13.0, 47.0] for eye-predominant effects, and 45.5 [25.0, 56.5] for brain-predominant effects (Figure 2b). Analysis of individual PCSI question scores showed that multiple symptoms were elevated in the brain-predominant effects group to a greater extent than the other two groups. Symptoms that were elevated (95% confidence intervals excluded zero) for the brain-predominant effects group included headache, light sensitivity, sound sensitivity, balance, dizziness, fatigue, drowsiness, slowed thinking, answering slowly, difficulty concentrating, mentally foggy, and visual problems (Figure S1).

**Figure 2:**
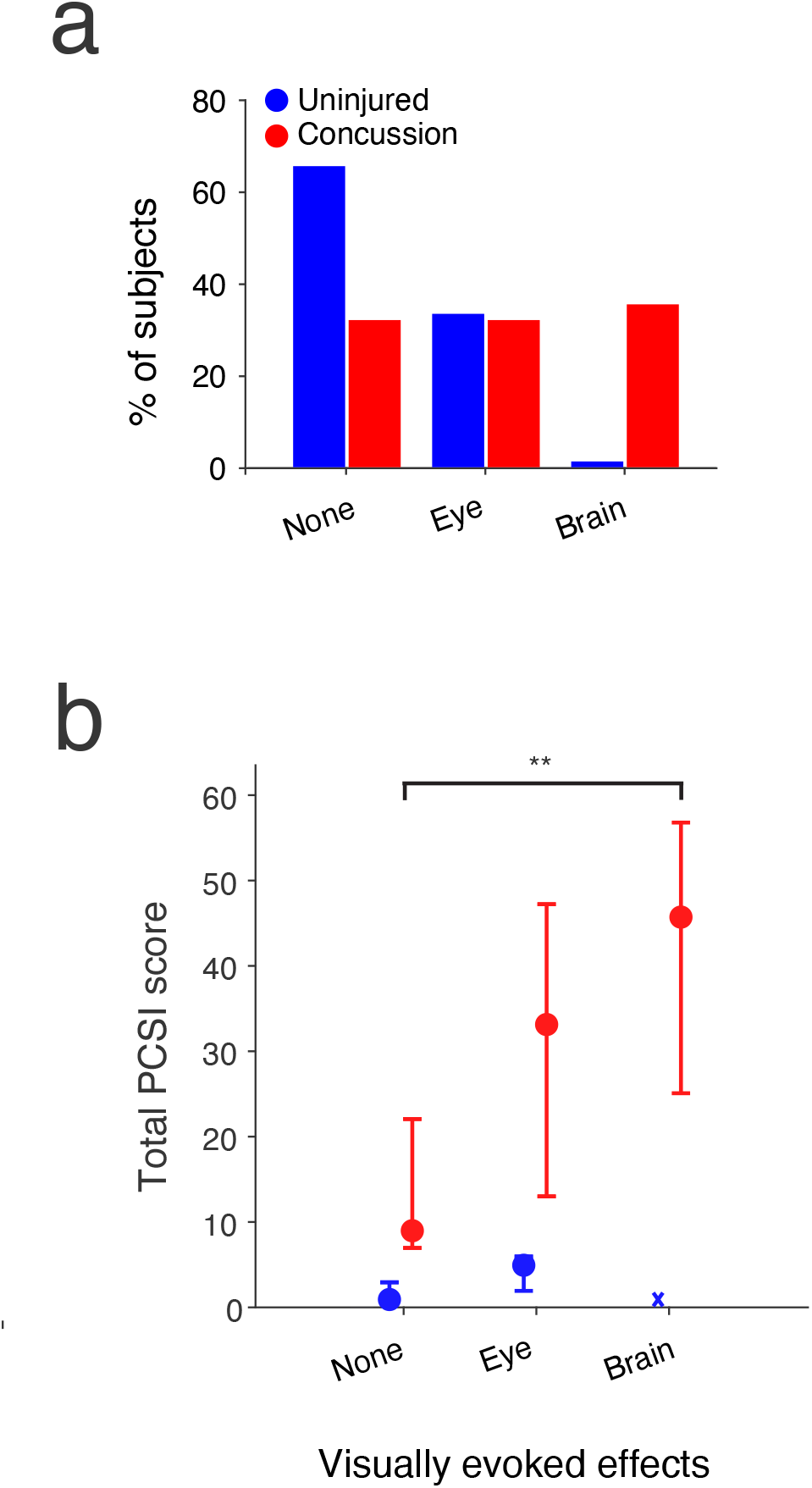
(a) Visually evoked effects categories of uninjured and concussed youth. (b) Total PCSI scores by visually evoked effects categories for uninjured and concussed youth.

There were no significant differences in migraine history, migraine family history, age, or biological sex across effect groups for uninjured or concussion participants (Table 2). Concussed participants in the no effects group were more likely to have had a history of concussion than those in the eye-predominant effects and brain-predominant effects groups (*χ*^2^ = 9.3, p = 0.01).

**Table 2:** Statistical comparison of visually evoked effects categories for uninjured and concussed youth.

|  | No effects | Eye effects | Brain effects | Stats |
| --- | --- | --- | --- | --- |
| **Participants (%)** | | | | $\chi^2 = 35.32, p = 2.1e^{-8}$ |
| Uninjured | 53 (65.4%) | 27 (33.3%) | 1 (1.2%) | -- |
| Concussion | 27 (32.1%) | 27 (32.1%) | 30 (35.7%) | -- |
| **Female/Male (% Female)** |  |  |  | -- |
| Uninjured | F 28, M 25 (52.8%) | F 14, M 13 (51.9%) | F 0, M 1 (0.0%) | $\chi^2 = 1.08, p = 0.58$ |
| Concussion | F 14, M 13 (51.9%) | F 17, M 10 (63.0%) | F 19, M 11 (63.3%) | $\chi^2 = 0.96, p = 0.61$ |
| **Median Age [95%CI]** |  |  |  |  |
| Uninjured | 16 [15, 17] | 16.0 [15, 17] | 13 [--] | $\chi^2 = 2.40, p = 0.30$ |
| Concussion | 16 [15, 16] | 16.0 [15, 16] | 16 [15, 16] | $\chi^2 = 0.64, p = 0.73$ |
| **History of migraine (%)** |  |  |  |  |
| Uninjured | 3 (5.6%) | 2 (7.4%) | 0 (0.0%) | $\chi^2 = 0.23, p = 0.89$ |
| Concussion | 2 (7.4%) | 5 (18.5%) | 4 (13.3%) | $\chi^2 = 0.85, p = 0.65$ |
| **Family history of migraine (%)** |  |  |  |  |
| Uninjured | 6 (11.3%) | 2 (7.4%) | 0 (0.0%) | $\chi^2 = 0.85, p = 0.72$ |
| Concussion | 9 (33.3%) | 8 (29.6%) | 12 (40.0%) | $\chi^2 = 0.23, p = 0.89$ |
| **History of concussion (%)** |  |  |  |  |
| Uninjured | 12 (22.6%) | 9 (33.3%) | 1 (100.0%) | $\chi^2 = 3.7, p = 0.16$ |
| Concussion | 18 (66.7%) | 11 (40.7%) | 8 (26.7%) | $\chi^2 = 9.3, p = 0.01$ |
| **Median Days post-injury [95% CI]** |  |  |  |  |
| Uninjured | -- | -- | -- | -- |
| Concussion | 14 [9, 18] | 11 [8, 16] | 13 [7, 16] | $\chi^2 = 0.66, p = 0.72$ |
| **Median total PCSI score [95%CI]** |  |  |  |  |
| Uninjured | 1.0 [0.0, 2.0] | 5.0 [2.0, 6.0] | 1 [1.0, 1.0] | $\chi^2 = 5.70, p = 0.06$ |
| Concussion | 0 [7.0, 22.0] | 33.0 [13.0, 47.0] | 45.5 [25.0, 56.5] | $\chi^2 = 15.39, p = 4.5e^{-4}$ |

### Concussion and brain-predominant visually evoked effects are associated with worse performance on saccadic, VOR, and VMS repetitions

The number of VVE repetitions participants were able to complete were compared between the effect categories, and between uninjured and concussion groups (Figure 3). Two-way ANOVA with interaction revealed significant differences between uninjured and concussion groups for horizontal and vertical saccades, horizontal and vertical VOR, and VMS, but no differences between concussion and uninjured participants in smooth pursuit repetitions or NPC (Table 3). Only horizontal VOR showed a significant effect of visually evoked groups, and there was no interaction between effect categories and uninjured versus concussion groups. However, multiple comparison analysis revealed that concussed participants with brain-predominant effects performed significantly worse compared to all other groups on horizontal saccades (p<0.01 for all comparisons), vertical saccades (p<0.001 for all comparisons), vertical VOR (p<0.02 for all comparisons), and VMS (p<0.01 for all comparisons). Overall, these findings show that participants who experienced brain-predominant effects had worse performance on multiple VVE tasks.

**Table 3:** Statistical analysis of VVE metrics by visually evoked effects categories and uninjured vs. concussion groups.

|  | Two-way ANOVA |  |  |
| --- | --- | --- | --- |
|  | Visually evoked effects | Uninjured v. concussion | Interaction |
| Smooth pursuits | F = 0.60, p = 0.55 | F = 0.34, p = 0.56 | F = 0.60, p = 0.55 |
| Near-point convergence | F = 0.38, p = 0.69 | F = 0.63, p = 0.43 | F = 0.10, p = 0.91 |
| Horizontal saccades | F = 1.06, p = 0.35 | F = 12.61, p = 5.0e<sup>-4</sup> | F = 0.74, p = 0.48 |
| Vertical saccades | F = 2.54, p = 0.08 | F = 13.99, p = 3.0e<sup>-4</sup> | F = 2.01, p = 0.14 |
| Horizontal VOR | F = 3.65, p = 0.03 | F = 23.41, p = 3.1e<sup>-6</sup> | F = 2.78, p = 0.07 |
| Vertical VOR | F = 1.90, p = 0.15 | F = 21.00, p = 9.2e<sup>-6</sup> | F = 1.90, p = 0.15 |
| VMS | F = 0.58, p = 0.56 | F = 4.36, p = 0.04 | F = 0.58, p = 0.56 |

**Figure 3:**
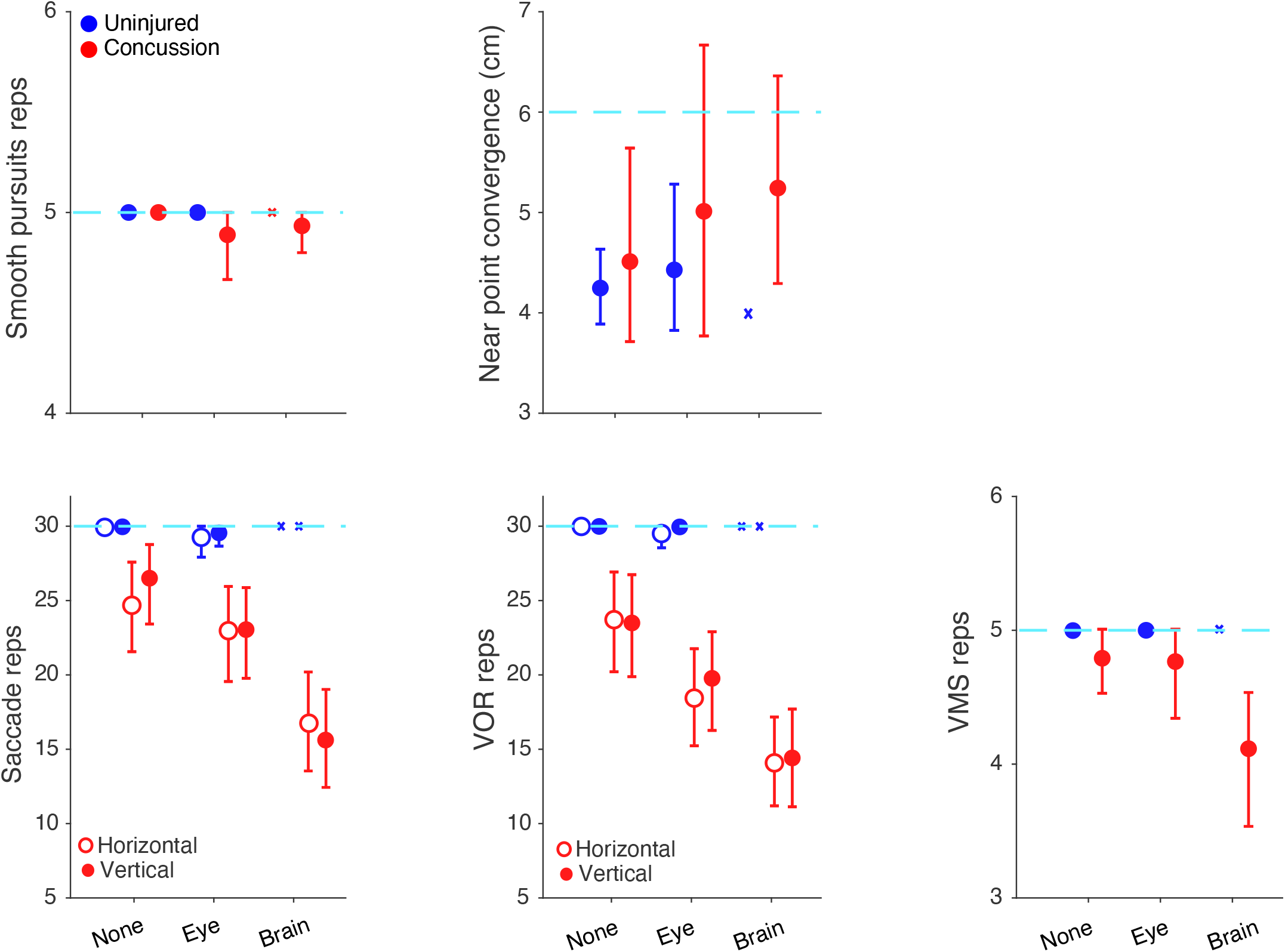
VVE metrics by visually evoked effects categories.

## DISCUSSION

Our study identified a relationship between the experience of negative somatic effects evoked by flickering light, the presence of concussion, and other characteristics of concussive injury, among a cohort of concussed and uninjured adolescents. We found that participants exposed to patterned flickering light differ along two primary axes: the presence or absence of evoked effects, and the degree to which these effects reflect predominantly eye-localized or more constitutional “brain” effects. Unsurprisingly, uninjured participants were most likely to experience no effects. Eye-related effects were present in similar proportion in the uninjured (33.3%) and concussed (32.1%) youth. This suggests that such effects may not be directly related to concussion, but rather evidence of a common premorbid sensitivity. This is consistent with the finding that 24% of uninjured neurologically-normal children presenting to an emergency department were found to have abnormalities in visio-vestibular testing^23^. Brain-predominant effects were uniquely associated with concussion. These effects were also associated with a greater symptom burden including multiple physical, fatigue, and cognitive concussion symptoms. Those with brain-predominant effects also showed the greatest deficits on visio-vestibular tasks that produced rapidly changing visual and/or vestibular input, similar to a flickering visual stimulus. Interestingly, the specificity and sensitivity of saccadic and VOR repetitions for identifying concussion has been optimized at 20 repetitions^7^. We measured 30 repetitions for both tasks and found that only the brain-predominant group consistently had a mean repetition count of less than 20 before symptoms were provoked.

While there is some inherent ambiguity in separating visually evoked effects into categories because their origins likely involve complex and intertwined neural pathways, the clusters are intriguing. Eye pain, tearing, and conjunctival injection have been associated with trigemino-vascular and trigemino-autonomic reflexes that involve trigeminal nerve afferents and brainstem nuclei^8^. Visually evoked eye effects may not require further activation beyond the reflex arch. The presence of multiple symptoms across different domains and impaired performance on multiple VVE tasks supports the idea that visually evoked brain effects reflect broader neurologic dysfunction. Indeed, constitutional somatic sensations including headache, dizziness, and nausea involve multiple cortical and subcortical brain regions^31–33^ and thus likely require contributions from both bottom-up and top-down processes. Communication between trigeminal reflexes and thalamocortical circuitry is bidirectional: activation of trigeminal afferents provides input to thalamocortical circuits^8^, which has been used to explain the multiple sensory and cognitive symptoms accompanying migrainous headache^34^. Thalamocortical circuits can also promote activation of trigeminal nociceptive pathways through disruption of inhibitory descending pain modulation and cortical spreading depression, which are also both proposed to underly post-traumatic headache^35–38^.

Activation of trigeminal nociceptive pathways has been extensively studied in migraine pathogenesis, and may help explain why there is symptom overlap between migraine and concussion^26,27,39^. Indeed, multiple studies have proposed a migrainous concussion phenotype that has been associated with high symptom burden and prolonged recovery ^40–44^. The brain predominant group in our study reported elevation of headache, and light and sound sensitivity, which are cardinal features of migraine^45^. Additionally, fatigue, dizziness, and cognitive complaints are common in individuals with chronic migraine^46,47^. We did not find a significant difference in distribution of visually evoked effects categories for youth with a family or personal history of migraine. However, the higher percentage of participants with a family history of migraine in the concussed participant group may suggest these individuals were predisposed to having symptoms necessitating concussion evaluation. Alternatively, high symptom burden and impairment on VVE in the brain effects category may simply reflect more widespread neural dysfunction as opposed to a particular concussion phenotype. Indeed, one of the most consistent predictors of prolonged recovery is overall symptom burden^28,48^, and high correlation across all symptoms is present even in studies that identified concussion phenotypes^44,49^. Further research is needed to elucidate the relationship between migraine, concussion, and visually evoked effects.

### Conclusions and future directions

This study demonstrates an intriguing relationship between brain-predominant visually evoked effects, concussion symptom burden, and reduced visio-vestibular performance. Though the study was limited by retrospective design, these strong associations warrant further exploration. The presence of visually evoked effects has great potential as an inexpensive, easily implemented, real-time psychophysical metric that could be used on the sideline of a sports field, the emergency department, or outpatient setting to supplement other concussion assessments. Prospective longitudinal studies are needed to validate this tool. It is not known if visually evoked effects are a prognostic indicator of prolonged recovery, or if they can predict treatment response to different therapies. Additionally, it should be determined if other neurologic conditions like chronic migraine are associated with visually evoked brain effects. It may be that visually evoked brain effects are a biomarker of broad activation of the trigeminal nociceptive pathway across a range of neurologic conditions.

## Data Availability

All data reported in the present study are available upon request to the authors.

https://github.com/pattersongentilelab/visuallyEvokedEffects.git

## Acknowledgements

The authors would like to thank Daniele Fedonni, Fairuz Mohammed, Olivia Podolak, and Anne Mozel who were instrumental in data collection and organization that made this work possible.

## Abbreviations

MCA: Multiple correspondence analysis
NPC: Near point convergence
PCSI: Post-concussion symptom inventory TBI Traumatic brain injury
VMS: Visual motion sensitivity
VOR: Vestibular-ocular reflex
VVE: Visio-vestibular examination

## FIGURE AND TABLE LEGENDS

**Table 1**: Demographics of uninjured and concussion groups. Kruskal Wallis *χ*^2^ test was used for statistical comparison.

**Figure 1:** (a) MCA of visually evoked effects. Individual effects are mapped onto this two-dimensional space. Dimension 1 represents the absence (negative direction) or presence (positive direction) of visually evoked effects, and dimension 2 divides eye effects (negative direction) and brain effects (positive direction). Mean MCA dimension scores are show for uninjured (blue) and concussed (red) youth. Error bars represent 95% confidence intervals by bootstrap analysis. (b) Dimension 1 and dimension 3 coefficients for individual uninjured (blue) and concussed (red) youth. Boxes represent visually evoked effects categories.

**Figure 2:** (a) Comparison of % of uninjured (blue) and concussed (red) youth by visually evoked effects category. (b) Median total PCSI values by visually evoked effects category for uninjured and concussion youth. Error bars represent 95% confidence intervals by bootstrap analysis. ‘X’ indicates the single uninjured participant in the brain-predominant effects category. Kruskal Wallis testing showed a significant difference of total PCSI score for concussion youth (*χ*^2^ = 15.4, p = 4.5e^-4^). Multiple comparisons using Tukey method found a significant difference in total PCSI scores between no effects and brain-predominant effects categories (**p = 3.0e^-4^).

**Table 2:** Comparison of visually evoked effects categories for uninjured and concussed groups. Kruskal Wallis *χ*^2^ test was used for statistical comparison.

**Figure 3:** Mean smooth pursuit repetitions, NPC, horizontal and vertical saccade repetitions, horizontal and vertical VOR repetitions, and VMS repetitions for uninjured (blue) and concussed (red) youth. Light blue dotted line represents target number of repetitions, or threshold for NPC insufficiency based on VVE criteria. Error bars represent 95% confidence intervals by bootstrap analysis. ‘X’ indicates the single uninjured participant in the brain-predominant effects category.

**Table 3:** ANOVA for visio-vestibular repetitions across visually evoked effects categories and ininjured versus concussion groups, with interaction.

