## Supplemental table 1 and figure 1 for "The relationship between visually evoked effects and concussion in youth"

| VISUALLY EVOKED SYMPTOM/ SIGN | VISUALLY EVOKED SYMPTOM CATEGORY: NONE Uninjured (n = 53) | NONE Concussion (n = 27) | EYE Uninjured (n = 27) | EYE Concussion (n = 27) | BRAIN Uninjured (n = 1) | BRAIN Concussion (n = 30) |
|---|---|---|---|---|---|---|
| Headache | 0 (0%) | 0 (0%) | 0 (0%) | 7 (26%) | 1 (100%) | 20 (67%) |
| Dizziness | 0 (0%) | 0 (0%) | 0 (0%) | 0 (0%) | 0 (0%) | 12 (40%) |
| Nausea | 0 (0%) | 0 (0%) | 0 (0%) | 0 (0%) | 0 (0%) | 8 (27%) |
| Eye reddening | 0 (0%) | 0 (0%) | 8 (30%) | 14 (52%) | 0 (0%) | 2 (7%) |
| Eye watering | 0 (0%) | 0 (0%) | 20 (74%) | 21 (78%) | 0 (0%) | 11 (37%) |
| Eye pain | 0 (0%) | 0 (0%) | 3 (11%) | 12 (44%) | 0 (0%) | 8 (27%) |
| Eye fatigue | 0 (0%) | 0 (0%) | 3 (11%) | 7 (26%) | 0 (0%) | 0 (0%) |

Table S1: Percentage of participants with individual visually evoked effects by uninjured and concussion groups, and visually evoked effects category.

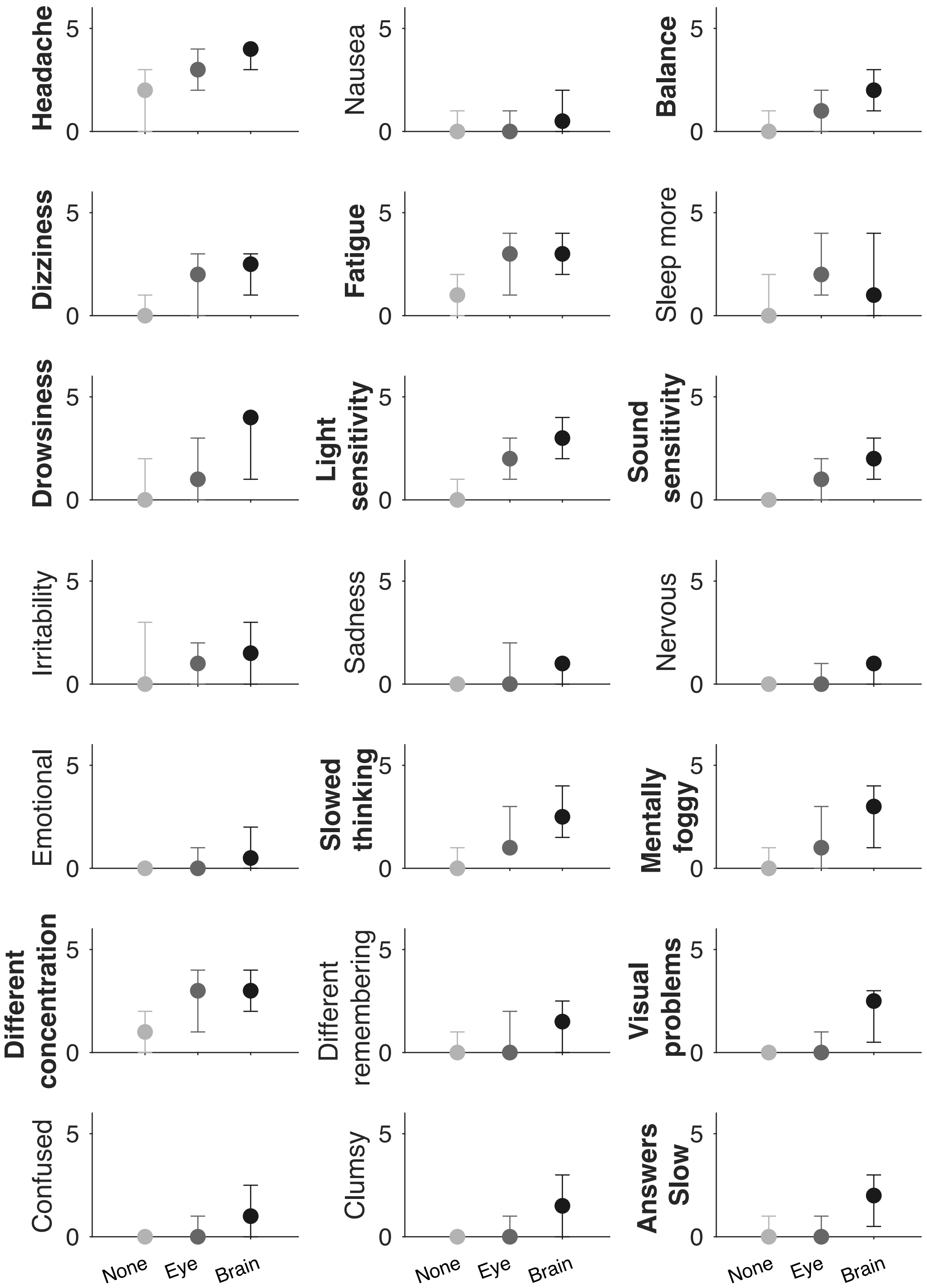


Figure S2: Individual median PCSI scores (0 - 6) for the visually evoked effects categories for concussed youth only. Error bars represent 95% confidence intervals by bootstrap analysis. Bolded y-axis labels indicate which symptoms were higher for the visually evoked brain effects category (95% CI did not include 0).
